# Rapid epidemic expansion of chikungunya virus-ECSA lineage in Paraguay

**DOI:** 10.1101/2023.04.16.23288635

**Authors:** Marta Giovanetti, Cynthia Vazquez, Mauricio Lima, Emerson Castro, Analia Rojas, Andrea Gomez de la Fuente, Carolina Aquino, Cesar Cantero, Fatima Fleitas, Juan Torales, Julio Barrios, Maria Jose Ortega, Maria Liz Gamarra, Shirley Villalba, Tania Alfonzo, Joilson Xavier, Talita Adelino, Hegger Fritsch, Felipe C. M. Iani, Glauco Carvalho Pereira, Carla de Oliveira, Gabriel Schuab, Evandra Strazza Rodrigues, Simone Kashima, Juliana Leite, Lionel Gresh, Leticia Franco, Houriiyah Tegally, Wesley C. Van Voorhis, Richard Lessels, Ana Maria Bispo de Filippis, Andrea Ojeda, Guillermo Sequera, Romeo Montoya, Edward C. Holmes, Tulio de Oliveira, Jairo Mendez Rico, José Lourenço, Vagner Fonseca, Luiz Carlos Junior Alcantara

## Abstract

The spread of vector-borne viruses, such as CHIKV, is a significant public health concern in the Americas, with over 120,000 cases and 51 deaths in 2023, of which 46 occurred in Paraguay. Using a suite of genomic, phylodynamic, and epidemiological techniques, we characterized the ongoing large CHIKV epidemic in Paraguay.

**Article Summary Line:** Genomic and epidemiological characterization of the ongoing Chikungunya virus epidemic in Paraguay

## Text

Chikungunya is a mosquito-borne disease caused by the chikungunya virus (CHIKV), a single-stranded positive-sense RNA virus belonging to the Togaviridae family (1), which is transmitted to humans through the bite of infected Aedes mosquitoes. Classically, it is an acute self-limiting illness characterized by fever and severe joint pain, although persistent or relapsing joint pain can occur (1). Atypical and severe manifestations (including meningoencephalitis) have been reported, and death is usually associated with older ages and other underlying diseases. Mother-to-child transmission of CHIKV does occur and neonatal disease can be severe, with neurological, myocardial, or haemorrhagic disease (1). CHIKV can be classified into four distinct genotypes: the West African; the East/Central/South African (ECSA); the Asian, and the Indian Ocean lineage (IOL) (2,3). The first imported case of CHIKV in Paraguay was detected in June 2014, in a patient from the Dominican Republic (4). Here, using a suite of on-site genomic monitoring, phylodynamic and epidemiological approaches we characterize the large-scale and ongoing 2022-2023 CHIKV epidemic in Paraguay.

### The Study

We partnered with the Pan-American Health Organization (PAHO) to perform on-site genomic surveillance at the Laboratorio Central de Salud Pública in Asunción, Paraguay. From March 11 to 17, 2023, a team of molecular biologists from Brazil and Paraguay worked with a set of selected samples (based on cycle threshold - Ct ≤35 and availability of epidemiological metadata, generating 179 viral genomes (deposited in GenBank, accession number OQ775394-OQ775567 and OQ567722-OQ567725). Sequencing was by Nanopore technology (5). With these data in hand, we estimated phylogenetic trees to explore the evolutionary and epidemiological relationship of CHIKV in Paraguay to those of other sequences of this viral genotype sampled globally. Accordingly, we retrieved 715 CHIKV-ECSA genome sequences with associated lineage date and country of collection from GenBank, collected up to March 30, 2023. The relevant methods are fully described in Supplementary Material.

Since the first autochthonous infections in Paraguay in 2015, CHIKV has been detected in the country every year (**Figure S1A**). Based on reported suspected CHIKV infections, the country has so far experienced four epidemic waves in 2015, 2016, 2018 and 2023, all associated with the summer months (**Figure S1A**). Between October 2, 2022, and April 10, 2023, a total of 118,179 suspected and confirmed infections have been already reported, including 3,510 hospitalized cases, and 46 deaths (4, 6). Neonates have accounted for 0.3% (n=162) of these cases and 8 deaths. In addition, 294 suspected cases of acute meningoencephalitis have been reported, 125 (43%) of which have been attributed to CHIKV (5, 6). While yearly minimum temperatures across Paraguay have remained stable over the past 40 years, we noted that mean and maximum yearly temperatures have been steadily increasing over this period, with the rapid and large resurgence of CHIKV in 2022 coinciding with the highest mean temperatures ever reported (**Figure 1A**). Before 2022, confirmed infections were restricted to the Central, Paraguarí, and Amambay districts, with the Central district dominating the reports (**Figure S1B**). After viral resurgence in 2022, confirmed infections have been reported in all districts (**Figure S1C**).

**Figure 1.**
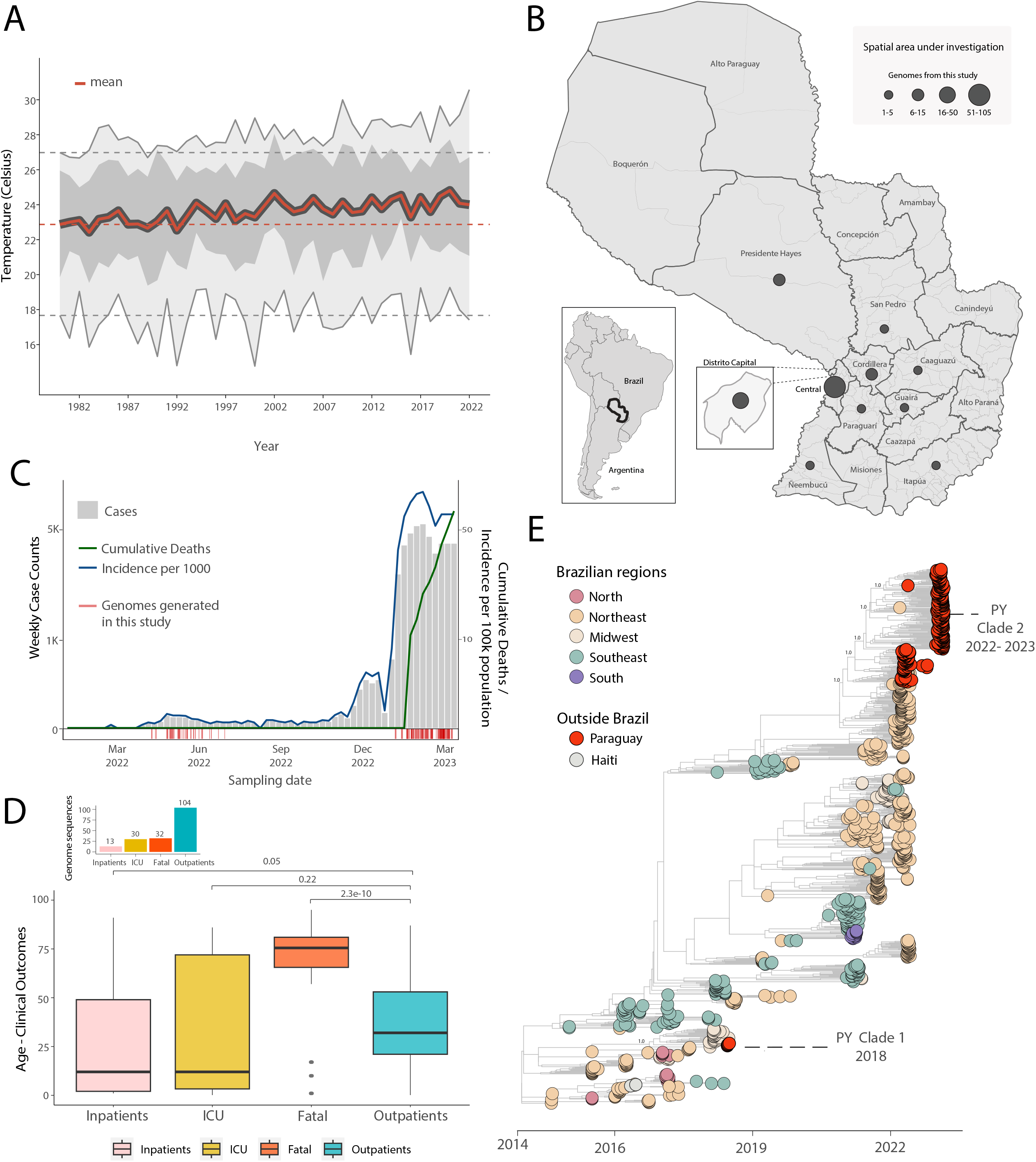
Spatial and temporal distribution of CHIKV cases in Paraguay. A) Temperature trends in Paraguay between 1981 and 2022. The yearly mean (red full line), yearly minimum and maximum (grey full lines), yearly 50% quantiles (dark grey area) and min, max and mean temperature in 1981 (dashed grey and red lines, respectively) are shown; B) Map of Paraguay showing the number of CHIKV genome sequences by departments. The size of the circles indicates the number of new genomes generated in this study; C) Weekly notified chikungunya cases (grey), incidence normalized per 100 K individuals (blue) and cumulative deaths (green) in 2022-2023 (until EW11), red bars at the bottom highlight the dates of sample collection of the genomes generated in this study; D) Boxplot of the patient’s (representing the study population) age and clinical outcomes value distribution. The Kruskal-Wallis non-parametric approach was used to determine the strength of association within the different clinical outcomes. E) Time-resolved maximum likelihood tree including the newly complete genome sequence from Paraguay (n=179) generated in this study combined with publicly available sequences (n=715) from GenBank collected up to March 30, 2023. Colors indicate geographic location of sampling. Support for branching structure is shown by bootstrap values at key nodes.

A total of 179 RT-qPCR positive samples for CHIKV were screened. All contained sufficient DNA (≥ 2 ng/µL) to proceed to library preparation, and their PCR Ct had a mean value of 21 (range: 9 to 34) (**Table S1**). The samples had a good spatial representation of southern Paraguay (10 out of 17 districts) (**Figure 1B**) including several of the districts with highest historical counts of CHIKV infections (**Figure S1B-D**) and captured both the out- and in-season periods of transmission (autumn and early winter 2022 and summer 2023, **Figure 1C**). Analyzing sample sequence coverage versus Ct revealed an average coverage of 94% among samples, and a Ct of 28 below which average coverage of >=90% was achieved (**Figure S2**). Most genomes (87%) were obtained from serum samples, and the rest from cerebrospinal fluid (CSF), while 54% were from females, and the mean age was 41 (range 26 days to 95) (**Table S1**). Most genomes were from CHIKV infection outcomes defined as outpatients (58%), followed by fatal (18%), intensive care unit (ICU, 17%) and inpatient (7%) infections (**Figure 1D** and **Supplementary Material** for definitions). Compared with an outpatient outcome, there was a clear association of fatal outcomes with older age-groups (**Figure 1D**). The same comparison with outcomes requiring medical attention (ICU, inpatients) was not statistically significant (**Figure 1D**). This latter observation contrasted the common notion that CHIKV symptomatic infections are more frequent in older age-groups (7).

To determine the dynamics of the CHIKV-ECSA in Paraguay, we performed phylodynamic analysis of a data set comprising 715 available representative genomes combined with the viruses sequenced in this study (n=179, collected between 06 April 2022 to 10 March 2023) (**Figure 1E**). A date-stamped phylogeny indicated that all the novel isolates formed a single large well-supported monophyletic group – denoted Paraguay clade 2 (PY2) - within the CHIKV-ECSA American clade. This strongly suggests that the 2022-2023 epidemic was not related to Brazilian cross-border transmission as in the past (8) (**Figure 1E, Clade PY I**), but was more likely the result of continual transmission within Paraguay over a period of 11 months of a viral strain that was introduced in the region in early 2022 (**Figure 1E, Clade 2, Figure 2C**).

**Figure 2.**
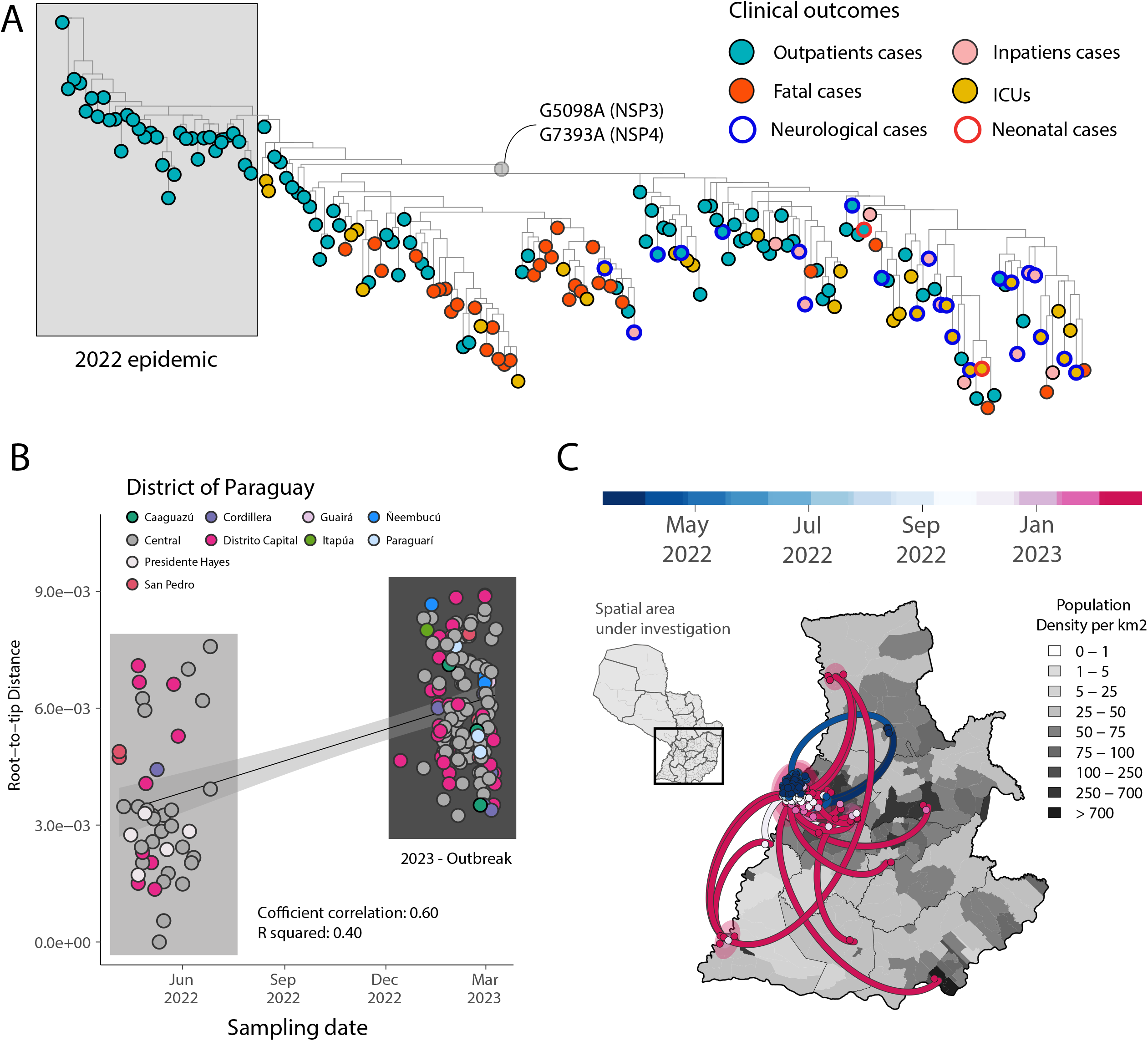
Expansion of the CHIKV-ECSA epidemic in Paraguay. A) Maximum Likelihood (ML) phylogeny constructed using n=174 CHIKV genome sequences from the PY clade 2 associated with the 2022-2023 epidemic. The tips were colored based on the clinical outcomes. Neurological and neonatal cases have been highlighted in the tree using blue and red borders, respectively. The length of a branch indicates the amount of sequence evolution; B) Regression of root-to-tip genetic distances and sampling dates estimated using TempEst v.1.5.3, buffers (shaded area) representing 90% confidence intervals. Colors indicate geographic location of sampling; C) Spatiotemporal reconstruction of the spread of CHIKV-ECSA in Paraguay. Circles represent nodes of the maximum clade credibility phylogeny, colored according to their inferred time of occurrence (scale shown). Shaded areas represent the 80% highest posterior density interval and depict the uncertainty of the phylogeographic estimates for each node. Solid curved lines denote the links between nodes and the directionality of movement. Differences in population density are shown on a grey-white scale.

To investigate the evolution of the PY clade 2 in more detail, we used a smaller data set (n=179) representing this virus clade in isolation. There was a relatively strong correlation between the sampling date and the root-to-tip genetic divergence in this data set (r^2^=0.40, coefficient correlation=0.60), indicating relatively clock-like virus evolution (**Figure 2B**). A phylogeographic analysis of PY2 allowed the reconstruction of viral movements among different districts in Paraguay (**Figure 2C**) and suggested a mean time of origin in late-March 2022 (95% highest posterior density (HPD): 25 March 2022 to 5 April 2022). Viruses from this clade spread multiple times from the Midwestern District (Distrito Capital and Central regions) towards the Southeast (Itapúa) and to the Midwest, as indicated by virus sequences from the Presidente Hayes and the Cordillera regions (**Figure 2C**). Transmission dynamics roughly followed patterns of population density, moving most often between the most populous urban localities (**Figure 1B, 2A, 2C**). Since it is recognized that not only non-synonymous, but also synonymous mutations can lead to changes in viral RNA (9, 10), affecting splicing, stability, translation or co-translational protein folding, additional studies will be necessary to determine the potential impact of these mutations on structure and function, and thus on viral pathogenesis and fitness.

## Conclusions

This study highlights the resurgence of CHIKV-ECSA in Paraguay in 2022-2023. Our findings provide evidence of lineage persistence in the country over a period of 11 months preceding resurgence and present the notable coincidence of virus resurgence alongside the highest mean temperatures ever recorded in Paraguay. These two factors, together with the presence of the vectors and a large proportion of the population susceptible to CHIKV, likely generated an ideal scenario for the observed fast and large CHIKV epidemic wave that started at the end of 2022. To date, the epidemic has been associated with a high frequency of severe symptoms and fatal outcomes in older age-groups. Given the association of the ongoing resurgence with a specific lineage of CHIKV-ECSA with two synonymous changes in nonstructural proteins (NSP3 and NSP4), and the uncertainty of how the ongoing epidemic will unfold, genomic surveillance should remain active to track its real-time evolution and spatial spread, in turn contributing to public health risk assessments in Paraguay and its South American neighbors.

## Ethics statement

This project was reviewed and approved by the Pan American Health Organization Ethics Review Committee (PAHOERC) (Ref. No. PAHO-2016-08-0029) and by the Paraguayan Ministry of Public Health and Social Welfare (MSPyBS/ S.G. no. 0944/18). The samples used in this study were de-identified residual samples from the routine diagnosis of arboviruses in the Paraguayan public health laboratory, which is part of the public network within the Paraguayan Ministry of Health.

## Biographical sketch

Dr. Giovanetti is a Visiting Researcher at the René Rachou Institute, Fiocruz Minas Gerais, Southeast Brazil and at the University Campus Bio-Medico in Rome, Italy. Her research focuses on investigating the patterns of gene flow in pathogen populations, focusing in phylogenetics and phylogeography as tools to recreate and understand the determinants of viral outbreaks and how this information can be translated into public policy recommendations.

## Data Availability

GenBank, accession number OQ775394-OQ775567 and OQ567722-OQ567725

## Acknowledgement

This work was supported by the PAHO Health Emergencies Department, by the National Institutes of Health USA grant U01 AI151698 for the United World Arbovirus Research Network (UWARN), FAPESP (2021/11944-6) and by Mercosur Structural Convergence Fund (FOCEM), Mercosur, FOCEM agreement N° 03/11 Project “Research, Education and Biotechnologies Applied to Health (COF 03/11). Data analysis support was provided by the Centre for Epidemic Response and Innovation (CERI) at Stellenbosch University supported by the Rockefeller Foundation. Authors would like to acknowledge the Global Consortium to Identify and Control Epidemics – CLIMADE, (T.O., L.C.J.A., E.C.H., J.L., M.G.) (https://climade.health/). The authors would like to express their gratitude to the Pan American Health Organization (PAHO/WHO), and in particular to Drs. Marcelo Korc, Alexander Rosewell, and Rodrigo Stabeli, for their significant contribution and assistance during this project. Because the journal had already achieved its limit for the number of writers, we were unable to include them on the paper as co-authors.

## Author contributions

Conception and design: M.G., C.V., J.L., J.M.R., L.C.J.A.; Investigations: M.G., C.V., M.L., E.C., A.R., A.G.d.l.F., C.A., C.C., F.F., J.T., J.B., M.J.O., M.L.G., S.V., T.A., J.X., T.A., H.F., F.C.M.I., C.d.O., G.S., E.S.R., S.K., J.L., L.G., L.F., H.T., R.L., A.M.B.d.F., A.O., G.S., R.M., M.K., E.C.H., T.d.O., J.M.R., J.L., V.F., and L.C.J.A; Data Analysis: M.G., H.T., J.L. and V.F.; Writing – Original: M.G., J.L., and L.C.J.A.; Revision: M.G., C.V., M.L., E.C., A.R., A.G.d.l.F., C.A., C.C., F.F., J.T., J.B., M.J.O., M.L.G., S.V., T.A., J.X., T.A., H.F., F.C.M.I., C.d.O., G.S., E.S.R., S.K., J.L., L.G., L.F., H.T., R.L., A.M.B.d.F., A.O., G.S., R.M., M.K., E.C.H., T.d.O., J.M.R., J.L., V.F., and L.C.J.A; Resources: C.V., J.M.R., and L.C.J.A.

## References

1. Schwartz O, Albert ML. Biology and pathogenesis of chikungunya virus. Nat Rev Microbiol. 2010;8(7):491–500. doi: 10.1038/nrmicro2368. PMID: 20551973.

2. de Oliveira EC, Fonseca V, Xavier J, Adelino T, Morales Claro I, Fabri A, Marques Macario E, Viniski AE, Campos Souza CL, Gomes da Costa ES, Soares de Sousa C, Guimarães Dias Duarte F, Correia de Medeiros A, Campelo de Albuquerque CF, Venancio Cunha R, Oliveira De Moura NF, Bispo de Filippis AM, Oliveira T, Lourenço J, de Abreu AL, Alcantara LCJ, Giovanetti M. Short report: Introduction of chikungunya virus ECSA genotype into the Brazilian Midwest and its dispersion through the Americas. PLoS Negl Trop Dis. 2021;16;15(4):e0009290.

3. Rico-Hesse, R. Molecular evolution and distribution of dengue viruses type 1 and 2 in nature. Virology. 1990; 174(2), 479–493.

4. Quick J, Grubaugh ND, Pullan ST, Claro IM, Smith AD, Gangavarapu K, et al. Multiplex PCR method for MinION and Illumina sequencing of Zika and other virus genomes directly from clin-ical samples. Nat Protoc. 2017;12:1261–76.

5. Pan-American Health Organization, CHIKV Weekly Report. PAHO. 2023. https://www3.paho.org/data/index.php/en/mnu-topics/chikv-en/550-chikv-weekly-en.html

6. Pan-American Health Organization, Disease Outbreak News. PAHO. 2023. https://www.who.int/emergencies/disease-outbreak-news/item/2023-DON448#:~:text=Paraguay%3A%20Between%202%20October%202022,hospitalized%20cases%20and%2046%20deaths.

7. Yoon IK, Alera MT, Lago CB, Tac-An IA, Villa D, Fernandez S, Thaisomboonsuk B, Klungthong C, Levy JW, Velasco JM, Roque VG Jr, Salje H, Macareo LR, Hermann LL, Nisalak A, Srikiatkhachorn A. High rate of subclinical chikungunya virus infection and association of neutralizing antibody with protection in a prospective cohort in the Philippines. PLoS Negl Trop Dis. 2015; 7;9(5):e0003764. doi: 10.1371/journal.pntd.0003764. PMID: 25951202; PMCID: PMC4423927.

8. Gräf T, Vazquez C, Giovanetti M, de Bruycker-Nogueira F, Fonseca V, Claro IM, de Jesus JG, Gómez A, Xavier J, de Mendonça MCL, Villalba S, Torales J, Gamarra ML, Thézé J, de Filippis AMB, Azevedo V, de Oliveira T, Franco L, de Albuquerque CFC, Irala S, Holmes EC, Méndez Rico JA, Alcantara LCJ. Epidemiologic History and Genetic Diversity Origins of Chikungunya and Dengue Viruses, Paraguay. Emerg Infect Dis. 2021; 27(5):1393–1404.

9. Faure, G., Ogurtsov, A.Y., Shabalina, S.A., and Koonin, E.V. Adaptation of mRNA structure to control protein folding. RNA Biol.2017; 14, 1649–1654.

10. Sharma Y, Miladi M, Dukare S, Boulay K, Caudron-Herger M, Groß M, Backofen R, Diederichs S. A pan-cancer analysis of synonymous mutations. Nat Commun. 2019; 12;10(1):2569.

