## Supplementary_Appendix for "Rapid epidemic expansion of chikungunya virus-ECSA lineage in Paraguay"

### Supplementary Material

#### Details on Methods & Data

##### Sample collection and whole genome sequencing

A total of 179 samples (n=156 serum, and n=23 cerebrospinal fluid - CSF) retrieved from patients presenting symptoms compatible with arboviral infection were collected by the Laboratorio Central de Salud Pública of Paraguay, in Asunción for molecular diagnosis. Samples were submitted first to nucleic acid extraction using the QIAmp Viral RNA Mini Kit (Qiagen) and then subjected to real-time reverse transcription PCR (RT-qPCR) for arbovirus detection (including ZIKV, CHIKV, ad DENV 1-4) (1-3). Genome sequencing was conducted using the Nanopore technology (4). Briefly, viral RNA was submitted to first-strand cDNA synthesis. Then, a multiplex tiling PCR was conducted using Q5 High Fidelity Hot-Start DNA Polymerase (New England Biolabs) and CHIKV sequencing primers (4). DNA library preparation was carried out using the Ligation Sequencing Kit and the Native Barcoding Kit (NBD104, Oxford Nanopore Technologies, Oxford, UK) (4). The final normalized sequencing library was loaded onto a R9.4 flow cell, and data were collected for 6 hours. FAST5 files were basecalled using Guppy and demultiplexing was performed using guppy software. Consensus sequences were obtained by hybrid assembly approach using Genome Detective (https://www.genomedetective.com/) (5). A total of 40,177 mapped reads were obtained, resulting in a sequencing mean depth >1,000X and a coverage of >94%, confirming CHIKV-ECSA genotype (**Table S1**).

Sequences were aligned using MAFFT (6) and edited using AliView (7). These datasets were assessed for the presence of phylogenetic signals by applying the likelihood mapping analysis implemented in the IQ-TREE2 software (8). A maximum likelihood phylogeny was reconstructed using IQ-TREE2 software under the HKY+G4 substitution model (8). We inferred time-scaled trees by using TreeTime (8). The presence of a temporal signal was evaluated in TempEst [(9),](https://sciwheel.com/work/citation?ids=1795582&pre=&suf=&sa=0&dbf=0) and time-scaled phylogenetic trees were inferred using the BEAST package (10). We employed a stringent model selection analysis using both path-sampling (PS) and steppingstone (SS) procedures to estimate the most appropriate molecular clock model for the Bayesian phylogenetic analysis (11). The uncorrelated relaxed molecular clock model was chosen for all datasets as indicated by estimating marginal likelihoods, also employing the codon based SRD06 model of nucleotide substitution and the nonparametric Bayesian Skyline coalescent model. To model the phylogenetic diffusion of detected 2022-2023 transmission clade we used a flexible relaxed random walk diffusion model (12,13) that accommodates branch-specific variation in rates of dispersal with a Cauchy distribution and a jitter window site of 0.01 (14,15). For each sequence, coordinates of latitude and longitude were attributed. MCMC analyses were performed in BEAST v1.10.4, running in duplicate for 50 million interactions and sampling every 10,000 steps in the chain. Convergence for each run was assessed in Tracer (effective sample size for all relevant model parameters >200). MCC trees for each run were summarized using TreeAnnotator after discarding the initial 10% as burn-in. Finally, we used the R package ‘seraphim’ version 1.0 (15) to extract and map spatiotemporal information embedded in the posterior trees.

Epidemiological data

Epidemiological data of weekly fatal, notified and laboratory confirmed cases CHIKV in Paraguay from 2013 to 2023 (Figure 1) were obtained and curated from the PAHO data repository for Chikungunya (17). Confirmed infections are defined as “a suspected or probable chikungunya case with a chikungunya test with positive result” (as stated on the PAHO platform).

Epidemiological data presented in Supplementary Figure S1 was provided by Dirección General de Vigilancia de la Salud del Ministerio de Paraguay (DGVS), including suspected, probable and confirmed CHIKV infections between 2015 and 2023 (17). Suspected infections are defined as any person with sudden onset of fever and arthralgia or disabling arthritis of sudden onset not explained by another medical condition. Probable infections are defined as any suspected case with a positive laboratory result for CHIKV (IgM ELISA) or any suspected case of CHIKV with an epidemiological link with a confirmed case. Confirmed infections are any suspected or probable case of CHIKV that includes real-time RT-PCR or viral isolation. When epidemiological data is presented, we aggregate suspected and probable infections into a single, suspected category.

Sample metadata

Samples were selected for sequencing based on the Ct value (≤35) and availability of epidemiological metadata, such as date of symptom onset, date of sample collection, sex, age, municipality of residence, symptoms, and disease classification (Table S1). Patients were classified based on their clinical outcomes in: Outpatient, Inpatient, intensive care unit (ICU), and fatal cases.

Temperature data

Monthly temperature data for Paraguay was extracted from Copernicus.eu satellite climate data (18). We summarized the temperature data by calculating the minimum, mean and maximum per year.

Generalized additive model of sample sequence coverage versus CT

We consider the sequencing coverage of each sample (between 0 and 1) as a probability that all genome sites are sequenced with success. For this, we augmented the dataset by counting the number of successful and unsuccessful events (sequencing of sites) per sample, from which we model a binomial based Generalised Additive Model (GAM). GAM was implemented using R v3.6.3 and the package mgcv v1.38.1 (19, 20). We included random effects for the clinical/infection outcome associated with each sample (“outcomes”) and for each sample independently (“ID”). The following code snippet summarizes this approach.

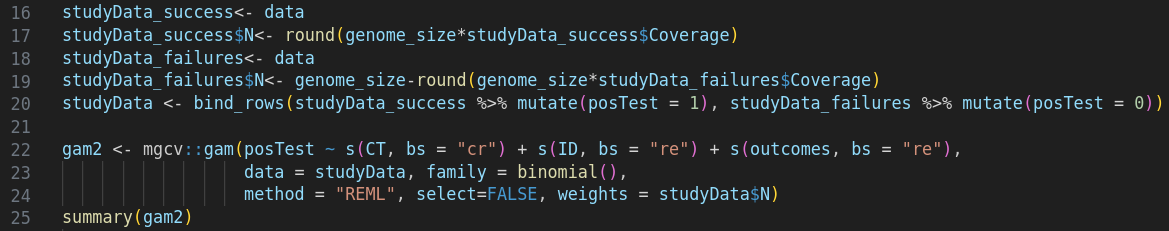

#### Supplementary Figures

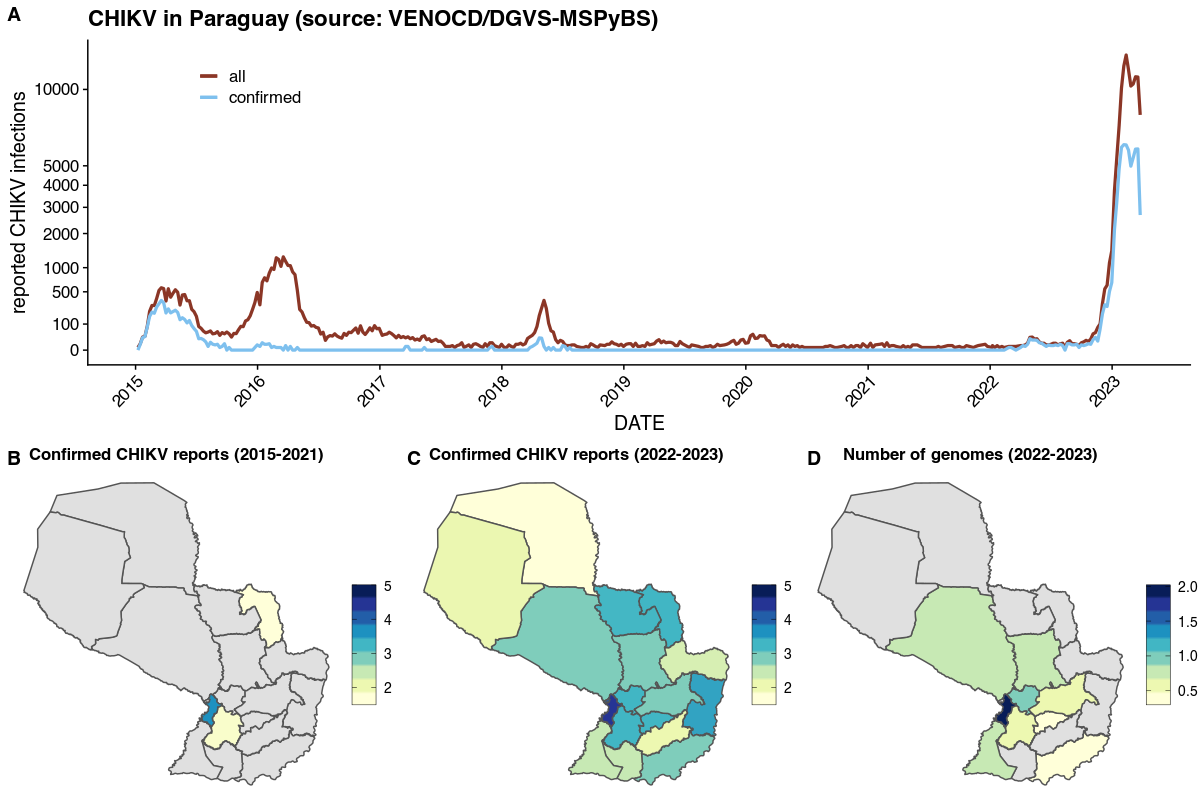

**Figure S1 - CHIKV reported infections and sequencing effort. (A)** Confirmed (in blue) and suspected plus confirmed (in red) CHIKV infections aggregated at the country level (Paraguay) as reported by Dirección General de Vigilancia de la Salud del Ministerio de Paraguay (DGVS) between 2015 and March 2023. **(B)** Spatial distribution of the sum of confirmed CHIKV infections per district in Paraguay between 2015 and 2021. **(C)** Spatial distribution of the sum of confirmed CHIKV infections per district in Paraguay between 2022 and 2023. **(D)** Spatial distribution of the total number of generated viral genomes in this study per district in Paraguay between 2022 and 2023. **(B-D)** Epidemiological data as reported by Dirección General de Vigilancia de la Salud del Ministerio de Paraguay. All color scales transformed by log10.

**Figure S2 - Generalized Additive Model of sample sequence coverage versus CT. (A)** Augmented fitted data as detailed in the supplementary text. **(B)** GAM predicted the probability of covering all genome sites with sequencing depending on the CT of each sample. Points are the samples, the red line is the mean predicted probability, the dark shaded area is the 50% CI and the light shaded area is the 95% CI.
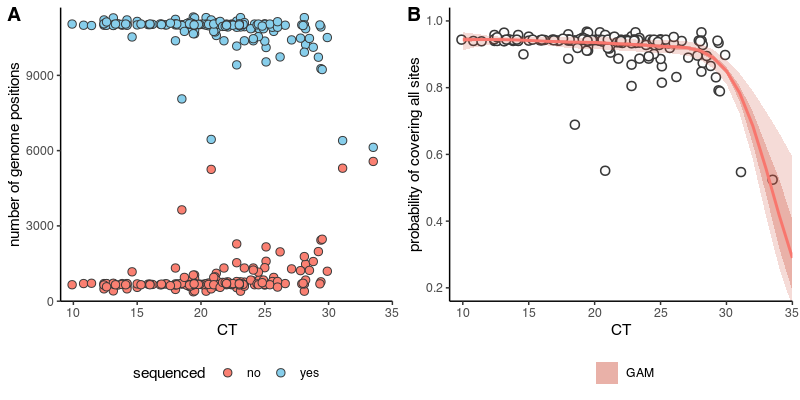

**References**

1. Lanciotti RS, Kosoy OL, Laven JJ, Velez JO, Lambert AJ, Johnson AJ, et al. Genetic and serologic properties of Zika virus associated with an epidemic, Yap State, Micronesia, 2007. Emerg Infect Dis. 2008;14:1232–9.
2. Santiago GA, Vergne E, Quiles Y, Cosme J, Vazquez J, Medina JF, Medina F, Colón C, Mar-golis H, Muñoz-Jordán JL. Analytical and clinical performance of the CDC real time RT-PCR as-say for detection and typing of dengue virus. PLoS Negl Trop Dis. 2013; 11;7(7):e2311.
3. Lanciotti RS, Kosoy OL, Laven JJ, Panella AJ, Velez JO, Lambert AJ, et al. Chikungunya vi-rus in US travelers returning from India, 2006. Emerg Infect Dis. 2007;13:764–7.
4. Quick J, Grubaugh ND, Pullan ST, Claro IM, Smith AD, Gangavarapu K, et al. Multiplex PCR method for MinION and Illumina sequencing of Zika and other virus genomes directly from clin-ical samples. Nat Protoc. 2017;12:1261–76.
5. Vilsker M, Moosa Y, Nooij S, Fonseca V, Ghysens Y, Dumon K, et al. Genome Detective: an auto-mated system for virus identification from high-throughput sequencing data. Bioinformatics. 2019; 1;35(5):871–3.
6. Katoh K, Rozewicki J, Yamada KD. MAFFT online service: multiple sequence alignment, interac-tive sequence choice and visualization. Brief Bioinform. 2019; 19;20(4):1160-1166.
7. Larsson A. AliView: a fast and lightweight alignment viewer and editor for large datasets. Bio-informatics. 2014; 30:3276–8.
8. Nguyen L-T, Schmidt HA, von Haeseler A, Minh BQ. IQ-TREE: a fast and effective stochastic al-gorithm for estimating maximum-likelihood phylogenies. Mol Biol Evol. 2015;32(1):268–74.
9. Rambaut A, Lam TT, Max Carvalho L, Pybus OG. Exploring the temporal structure of heter-ochronous sequences using TempEst (formerly Path-O-Gen). Virus Evol. 2016; 2:vew007.
10. Suchard MA, Lemey P, Baele G, Ayres DL, Drummond AJ, Rambaut A. Bayesian phylogenetic and phylodynamic data integration using BEAST 1.10. Virus Evol. 2018; 4:vey016.
11. Baele, G., Li, W. L., Drummond, A. J., Suchard, M. A. & Lemey, P. Accurate model selection of re-laxed molecular clocks in bayesian phylogenetics. Mol. Biol. Evol. 2013; 30, 239–243.
12. Lemey P, Rambaut A, Welch JJ, Suchard MA. Phylogeography takes a relaxed random walk in continuous space and time. Mol Biol Evol. 2010; 27:1877–1885.
13. Pybus OG, Suchard MA, Lemey P, Bernardin FJ, Rambaut A, Crawford FW, Gray RR, Arinamin-pathy N, Stramer SL, Busch MP, Delwart EL. Unifying the spatial epidemiology and molec-ular evolution of emerging epidemics. Proc Natl Acad Sci. 2012; 109:15066–15071.
14. Dellicour, S. et al. Relax, keep walking - a practical guide to continuous phylogeographic inference with BEAST. Mol. Biol. Evol. 2021; 38,3486–3493.
15. Dellicour, S., Rose, R., Faria, N. R., Lemey, P. & Pybus, O. G. SERAPHIM:studying environ-mental rasters and phylogenetically informed movements. Bioinformatics. 2016, 32, 3204–3206.
16. Pan-American Health Organization, CHIKV Weekly Report. PAHO. 2023.<https://www3.paho.org/data/index.php/en/mnu-topics/chikv-en/550-chikv-weekly-en.html>
17. Paraguayan Ministry of Health. DGVS, Dirección General de Vigilancia de la Salud del Paraguay, 2023.<https://dgvs.mspbs.gov.py/enfermedades/chikungunya/>
18. Essential climate variables for assessment of climate variability from 1979 to present. Copernicus Climate Data Store.<https://cds.climate.copernicus.eu/cdsapp#!/dataset/ecv-for-climate-change>
19. Pedersen EJ, Miller DL, Simpson GL, Ross N. Hierarchical generalized additive models in ecology: an introduction with mgcv. PeerJ. 2019;7: e6876.
20. The R Project for Statistical Computing. [cited 4 Apr 2023]. Available: [https://www.R-project.org/](https://www.r-project.org/)
